# Deciphering the dynamic clinical patterns in *SCN8A*-related disorders using real-world data

**DOI:** 10.1101/2024.10.21.24315870

**Authors:** Jan H. Magielski, Stacey Cohen, Michael C. Kaufman, Shridhar Parthasarathy, Julie Xian, Elise Brimble, Nasha Fitter, Francesca Furia, Elena Gardella, Rikke S. Møller, Ingo Helbig, Jillian L. McKee

## Abstract

**Background and Objectives:** *SCN8A*-related disorders encompass a range of neurodevelopmental and epilepsy phenotypes. However, despite representing one of the most common epilepsy-associated channelopathies, the longitudinal progression of its clinical features remains largely uncharacterized.

**Methods:** Here, we harmonized electronic medical record data of 82 individuals with *SCN8A*-related disorders. Clinical data was mapped to the standardized language of the Human Phenotype Ontology to reconstruct the natural history of *SCN8A*-related disorders in comparison to a cohort of 2,833 individuals with known or presumed genetic epilepsies.

**Results:** Compared to other known or presumed genetic epilepsies, those with *SCN8A*-related disorders had earlier generalized seizures, including a ten-fold risk for generalized-onset seizures at 9 months (*P* = 4.98 × 10^−17^, OR 10.65, CI 6.07-18.77) and >40-fold risk bilateral tonic-clonic seizures at 1 year (*P* = 1.76 × 10^−8^, OR 46.09, CI 10.72-275.01). Individuals carrying gain-of-function *SCN8A* variants had particularly high generalized seizure risk at 9 months (*P* = 0.005, OR 3.85, CI 1.35-11.55), as well as an increased risk for global developmental delay as early as 3 months (*P* = 0.002, OR 5.67, CI 1.74-20.23) when compared to the broader *SCN8A* cohort. Individuals with loss-of-function variants were more likely to experience absence seizures, most prominently at 4.25 years (*P* = 0.013, OR 32.71, CI 1.44-2193.51). Compared to the wider *SCN8A* cohort, individuals with the recurrent p.Arg850Gln variant were more likely to have infantile spasms at 6 months (*P* = 0.016, OR 12.02, CI 1.20-164.23), those with variants at the p.Arg1872Trp/Gln/Leu hotspot were more likely to have neonatal seizures (*P* = 0.025, OR 16.84, CI 0.99-291.58). Individuals with the recurrent p.Gly1475Arg variant were more likely to have active epilepsy after 5 years of age. Focal seizures in later childhood were more prominent in individuals with the recurrent p.Arg1617Gln variant.

**Discussion:** *SCN8A*-related disorders distinguish themselves from other genetic epilepsies by the frequent generalized-onset seizures in infancy, prominent early epileptic and developmental features in gain-of-function variant carriers, and unique seizure phenotypes in those with recurrent variants. Our study provides a longitudinal perspective on this genetic condition, paving the way for the future precision medicine approaches.

## Introduction

*SCN8A* encodes the voltage-gated sodium channel Na_v_1.6.^1^ Rare variants in this gene are an established cause of epilepsy with a broad spectrum of severity, ranging from self-limiting familial infantile epilepsies to developmental and epileptic encephalopathies.^2–7^ *SCN8A*-related disorders are typically characterized by early-onset refractory seizures and neurodevelopmental abnormalities, and disease-causing variants in *SCN8A* are estimated to explain a significant fraction of epileptic encephalopathies.^8^ Since the initial discovery in 2012, *SCN8A*-related disorders have received significant attention given the frequency of the condition and the potentially targetable disease mechanism. Initial genotype-phenotype correlation and functional biophysical studies have been performed, allowing for a baseline understanding how gain- and loss-of-function variation contribute to different phenotypes.^2^ However, the longitudinal trajectory of *SCN8A*-related disorders, which is pivotal for developing clinical trial readiness and establishing rational outcomes measures, remains largely unexplored.

*SCN8A*-related epilepsies have a unique place among the sodium channelopathies, both in terms of clinical presentation and the underlying biophysical etiology. Unlike Dravet syndrome due to loss-of-function (LOF) *SCN1A* variants, mutations in *SCN8A* predominantly result in gain-of-function (GOF) effects.^8^ Na_V_1.6 is mostly expressed at the nodes of Ranvier and the axon initial segment, which makes it a crucial protein in initiating and maintaining neuronal communication.^1,3,9,10^ GOF as the predominant mechanism of the disorder offers potential for precision medicine approaches. Anti-sense oligonucleotide therapies lowering the expression of *SCN8A* mRNA could thus be a potentially effective intervention, as shown in mouse models.^11,12^

To advance clinical trial readiness, comprehensive delineation of natural histories in addition to translational research effects is crucial.^13^ Electronic medical records (EMR) have been increasingly used to characterize various neurological conditions using real-world data.^14–16^ By combining these data resources with standardized phenotyping frameworks and dictionaries such as the Human Phenotype Ontology (HPO), it is feasible to standardize clinical data and maximize the capture of clinical information from the EMR.^17,18^ Through this approach, phenotypic patterns can be comprehensively characterized over time, allowing for an in-depth description of subgroups, genotype-phenotype correlations, and dynamic ranges of clinical features over time.^19–24^

Here, we characterized the phenotypic spectrum of *SCN8A*-related disorders in 82 individuals across 604 patient years. We assessed seizure trajectories, neurodevelopmental histories, and medication responses, while further delineating relevant clinical subgroups. We demonstrate that *SCN8A*-related disorders have unique longitudinal phenotypic signatures distinct from other epilepsies and neurodevelopmental disorders.

## Methods

### Identification of the cohort

In our study, we included 82 individuals with *SCN8A*-related disorders. The cohort consisted of 67 individuals from the Citizen Health and 20 individuals followed at a tertiary care pediatric institution (Children’s Hospital of Philadelphia). The EMR data for each patient were reviewed and data of duplicates between both cohorts—as determined by the same date of birth, *SCN8A* variant, and biological sex—were combined. Five individuals were detected to be present in both datasets. All variants were reviewed by a licensed genetic counselor, and only the ones that met the American College of Medical Genetics and Genomics (ACMG) criteria for likely pathogenicity or pathogenicity were included. All participants provided informed consent to participate; the study was conducted in accordance with the Declaration of Helsinki and the STROBE guidelines.

### Standardizing the phenotypic information

We employed the HPO to standardize the clinical data captured from the EMR, a standardized dictionary with more than 13,000 phenotypic terms commonly used in genetic studies and clinical diagnostics.^25–27^ All clinical features contained within the EMR were manually translated to HPO terms. Following the process of translation, the terms were “propagated”, i.e. combined with all higher-level clinical terms within the tree-like pattern of the ontology, as described previously.^22^ This allowed us to track clinical terms higher on the ontological tree, filling in gaps for frequencies of higher-level concepts that are commonly provided in raw EMR or clinical data.^14,15,23^

To compare neurological features over time, we selected relevant clinical terms by focusing on terms in the branching, tree-like pattern of the HPO dictionary at or below seizure (HP:0001250), neurodevelopmental delay (HP:0012758), and abnormal muscle tone (HP:0003808). This enabled comparison of frequencies of neurological features with a cohort of 2,833 individuals with epilepsy and neurodevelopmental disorders with presumed genetic basis, recruited on an ongoing basis since 2014.

Clinical data for these 2,833 individuals were extracted from the EMR and translated to the HPO dictionary with the use of Natural Language Processing tools as previously described.^28^ Individuals in the comparator cohort (Epilepsy Genetics Research Project, EGRP) with *SCN8A* disorders were excluded. Phenotypic data was grouped into 3-month age bins and Fisher’s tests were performed for each time bin and each clinical feature. Similar analyses were performed for several subgroups within the *SCN8A* cohort compared to the remainder of the *SCN8A* cohort, including (1) GOF variants, (2) LOF variants, and (3) recurrent variants. Only features that were present in at least two individuals in the subgroup of interest and in at least one individual in the comparator group were included in the analysis.

### Reconstructing seizure histories

We reconstructed seizure histories through retrospective manual review of the available clinical records from the local *SCN8A* cohort (EGRP) as well as seizure histories reconstructed from Citizen Health. For each individual, we recorded the presence or absence of seizures and the seizure types as described by HPO terms for every month of available EMR data. Additionally, we used an established seizure severity scale championed by the Epilepsy Foundation’s Epilepsy Learning Health System (ELHS) and Pediatric Epilepsy Learning Healthcare System^29,30^ to reconstruct monthly seizure frequencies as follows: no seizures (SF score = 0), monthly seizures (SF score = 1), weekly seizures (SF score = 2), daily seizures (SF score = 3), 2-5 daily seizures (SF score = 4), and more than 5 daily seizures (SF score = 5).^23,31^

### Anti-seizure medication retrieval and analysis

We retrieved anti-seizure medication (ASM) data from the EMR for each individual in the EGRP cohort, as previously described.^23^ In parallel, medication information was also extracted from the Citizen Health dataset. For each ASM in each individual, ages at initiation and discontinuation were recorded. Common seizure rescue medications were excluded.

A total of 68 individuals had clinical information for both seizure frequency history and medication data. To assess the comparative ASM effectiveness in these 68 individuals, we analyzed the change of SF scores during exposure to specific ASMs, as previously reported.^23,31^ For example, if an individual was prescribed oxcarbazepine between three months to four years, seizure frequency changes during this period were compared to changes in seizure frequency during intervals without oxcarbazepine. By performing Fisher’s exact test, medication effectiveness across three categories was analyzed: seizure frequency reduction, seizure freedom maintenance, and seizure frequency reduction or seizure freedom maintenance, as previously determined for *STXBP1* disorders.^23,24^ Only ASMs that were taken by at least ten individuals were included in the comparative effectiveness analysis.

### Statistical analysis

Statistical analyses were conducted using the R Statistical Package.^32^ We used Fisher’s exact test to assess the significance of the clinical feature associations and medication comparative effectiveness analyses. To correct for multiple testing, the false discovery rate (FDR) was set for 5%. The findings that did not maintain significance after the FDR correction are still reported with their odds ratios (OR) and confidence intervals (CI).

### Data availability

Primary data for the analysis contain sensitive health information and cannot be made publicly viewable, but they are available in a de-identified form upon request from the corresponding author.

## Results

### *SCN8A*-related disorders encompass a spectrum of biophysical and phenotypic characteristics

We analyzed longitudinal clinical data from 82 individuals across 604 patient years, including a total of 131,524 clinical terms and 1,692 unique terms (**Figure 1**). Our cohort included 28 individuals with established GOF *SCN8A* variants and 6 with established LOF (including protein-truncating and frameshift mutations) variants (**Supplementary Table 1**). We identified six recurrent variants, including p.Arg1872Trp/Gln/Leu (*n*=6), p.Arg1617Gln (*n*=6), p.Arg850Gln (*n*=5), p.Gly1475Arg (*n*=4), p.Asn1877Ser (*n*=4), and p.Ile240Val/Leu (*n*=2).

**Figure 1.**
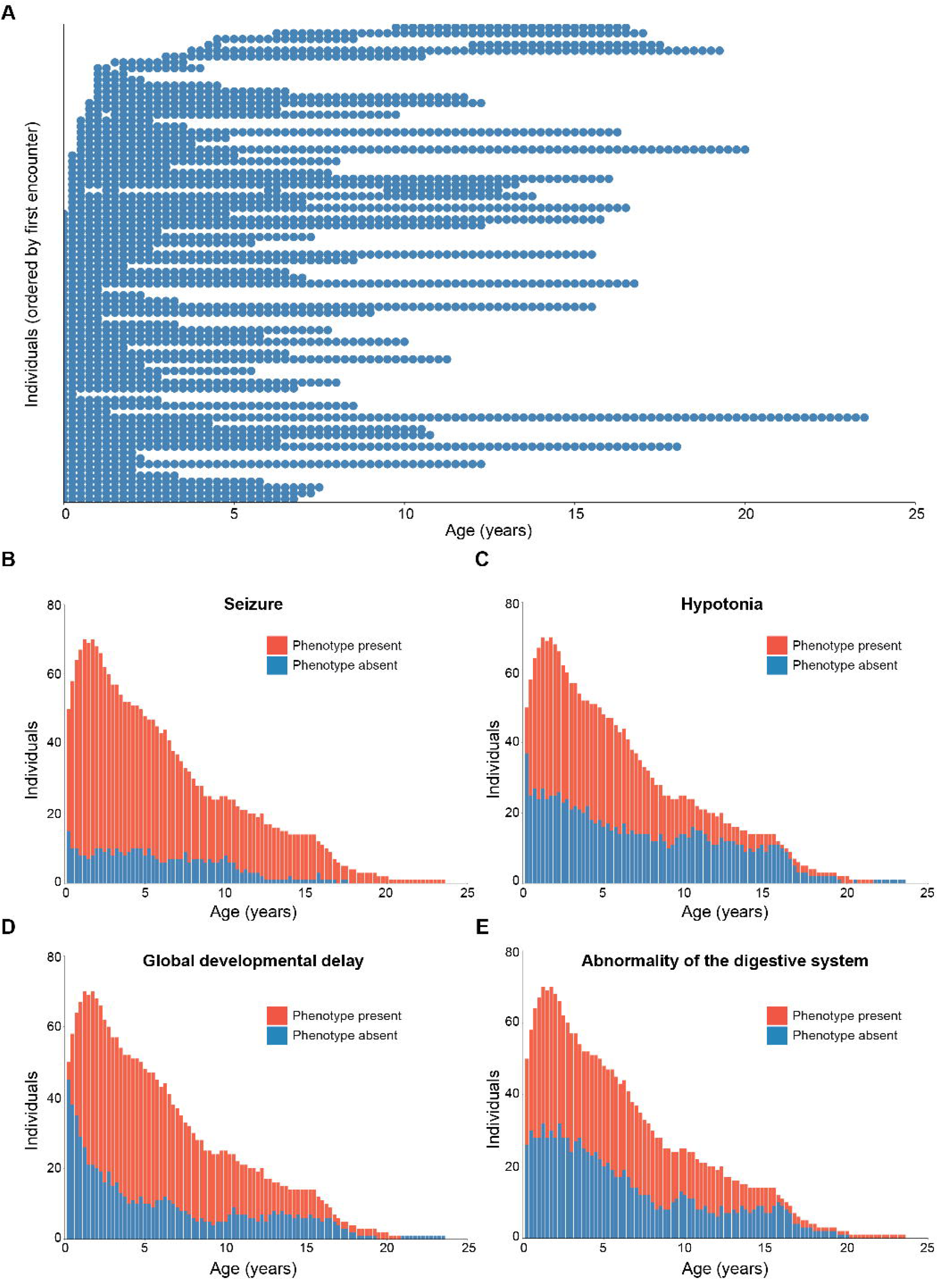
Longitudinal phenotypes. All encounters of 82 individuals with *SCN8A*-related disorders across 131,524 HPO terms and 604 patient-years (**A**). The longitudinal phenotype of seizure (**B**), hypotonia (**C**), global developmental delay (**D**), abnormality of the digestive system (**E**). The red color represents the presence of the phenotype, and the blue color its absence.

Common clinical traits in the cohort of 82 individuals with *SCN8A*-related disorders included seizures (*f* = 0.96), neurodevelopmental delays (*f* = 0.95), movement abnormalities (*f* = 0.95), hypotonia (*f* = 0.91), and behavioral abnormality (*f* = 0.90). For the GOF subgroup (*n* = 28), the most common clinical features included generalized-onset seizures (*f* = 1), EEG abnormalities (*f* = 0.96), abnormal muscle tone (*f* = 0.96), neurodevelopmental delay (*f* = 0.96), abnormality of the digestive system (*f* = 0.96), and hypotonia (*f* = 0.93). In the LOF subgroup (*n* = 6), seizures were seen less frequently (*f* = 0.67), with global developmental delay (*f* = 1), behavioral abnormality (*f* = 1), and hypotonia (*f* = 0.83) as the most common clinical features (**Table 1**).

**Table 1.** Overview of the most common phenotypes.

| All individuals ($n = 82$) | |
| --- | --- |
| Phenotype | Frequency |
| Abnormal muscle tone (HP:0003808) | 0.98 |
| Seizure (HP:0001250) | 0.96 |
| Neurodevelopmental delay (HP:0012758) | 0.95 |
| Abnormality of movement (HP:0100022) | 0.95 |
| Hypotonia (HP:0001252) | 0.91 |
| Behavioral abnormality (HP:0000708) | 0.90 |
| EEG abnormality (HP:0002353) | 0.90 |
| Abnormality of the digestive system (HP:0025031) | 0.89 |
| Generalized-onset seizure (HP:0002197) | 0.88 |
| Sleep disturbance (HP:0002360) | 0.73 |
| GOF ($n = 28$) | |
| Seizure (HP:0001250) | 1 |
| Generalized-onset seizure (HP:0002197) | 1 |
| EEG abnormality (HP:0002353) | 0.96 |
| Abnormal muscle tone (HP:0003808) | 0.96 |
| Neurodevelopmental delay (HP:0012758) | 0.96 |
| Hypotonia (HP:0001252) | 0.93 |
| Behavioral abnormality (HP:0000708) | 0.89 |
| Sleep disturbance (HP:0002360) | 0.86 |
| Motor seizure (HP:0020219) | 0.86 |
| Abnormality of the skeletal system (HP:0000924) | 0.82 |
| LOF ($n = 6$) | |
| Neurodevelopmental delay (HP:0012758) | 1 |
| Behavioral abnormality (HP:0000708) | 1 |
| Abnormal central motor function (HP:0011442) | 1 |
| Abnormal eye physiology (HP:0012373) | 1 |
| Hypotonia (HP:0001252) | 0.83 |
| Abnormality of movement (HP:0100022) | 0.83 |
| Anxiety (HP:0000739) | 0.67 |
| Intellectual disability (HP:0001249) | 0.67 |
| Seizure (HP:0001250) | 0.67 |
| EEG abnormality (HP:0002353) | 0.67 |

### Longitudinal phenotypes distinguish *SCN8A* from other epilepsies

By comparing the longitudinal phenotypes of individuals with *SCN8A*-related disorders to those of individuals with other epilepsies and neurodevelopmental disorders with a known or presumed genetic basis, we identified age-dependent phenotypic features that were more likely to occur in individuals with *SCN8A*-related disorders (**Figure 2**). Those carrying disease-causing *SCN8A* variants had a significantly higher burden of seizures, neurodevelopmental features, and motor abnormalities that were more prominent in the first five years of life.

**Figure 2.**
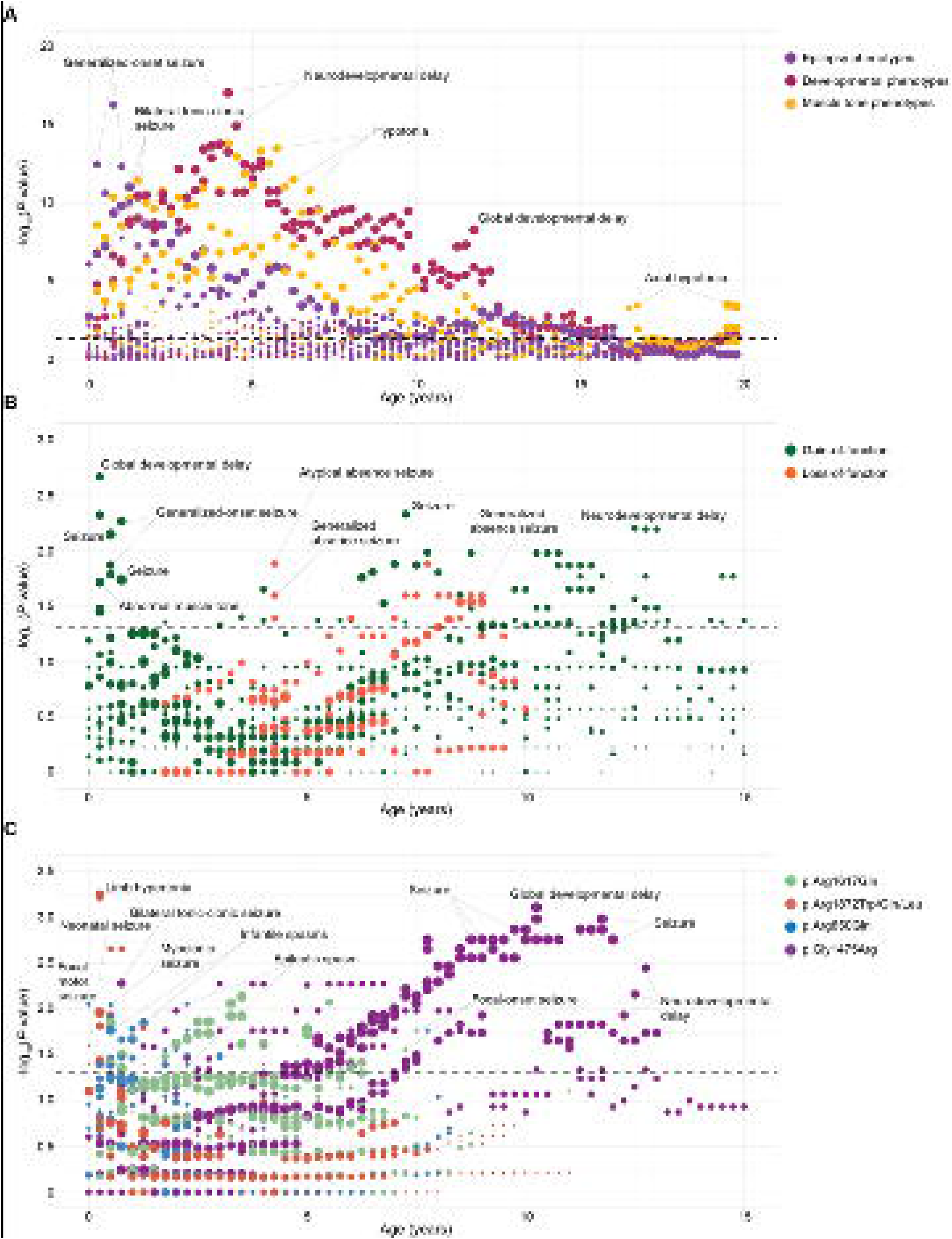
Longitudinal phenotypic comparison of *SCN8A*-related disorders. Comparison of *SCN8A*-related disorder phenotypes to other epilepsies (**A**). Comparison of the *SCN8A* GOF and *SCN8A* LOF variant carriers’ phenotypes to the remainder of the SCN8A cohort (**B**). Comparison of the *SCN8A* recurrent variant carriers’ phenotypes to the remainder of the *SCN8A* cohort (**C**). The data was binned into 3-month increments. The dot size on the plots reflects the frequency of a given phenotype. The dashed line represents the post-FDR significance threshold (**A**) or the nominal significance threshold (**B**, **C**).

Generalized-onset seizure was the most significant association within the epilepsy phenotypes, particularly at the age of three months (*P* = 3.32 × 10^−13^, OR 10.39, CI 5.51-19.55), nine months (*P* = 4.98 × 10^−17^, OR 10.65, CI 6.07-18.77), and one year (*P* = 4.52 × 10^−13^, OR 7.51, CI 4.34-12.95). Overall, the *SCN8A* cohort was more likely to experience any type of seizure across most time points, consecutively from 0 to 8.5 years, but also at later ages (10.75, 11.25-13.75, and 14-14.75 years). Other seizure types significant after correction for multiple testing included bilateral tonic-clonic seizures with focal onset, generalized tonic seizures, generalized-onset motor seizures, multifocal seizures, and focal clonic seizure (**Supplementary Table 2**). While the *SCN8A* cohort was not less likely to experience any seizure type following the FDR correction, the nominally significant associations included epileptic spasm in the first three months of life (*P* = 0.038, OR 0.34, CI 0.09-0.95), febrile seizure from 3 (*P* = 0.026, OR 0.24, CI 0.03-0.92) to 3.5 (*P* = 0.025, OR 0.24, CI 0.03-0.92) years of age, motor seizure from 3 (*P* = 0.023, OR 0.39, CI 0.14-0.92) to 3.75 (*P* = 0.038, OR 0.39, CI 0.12-0.99) years and from 4.5 (*P* = 0.030, OR 0.33, CI 0.09-0.92) to 5 (*P* = 0.029, OR 0.33, CI 0.08-0.91) years of age, and status epilepticus at 2.75 years of age (*P* = 0.038, OR 0.25, CI 0.03-0.98).

Among the various neurodevelopmental features, neurodevelopmental delay was the most prominent symptom associated with *SCN8A*-related disorders, including significance after multiple testing correction. The strongest association was observed at 4.25 years of age (*P* = 8.33 × 10^−18^, OR 23.48, CI 8.51-90.34), indicating a more than 20-fold increase in enrichment for neurodevelopmental features in *SCN8A*-related disorders compared to other epilepsies. This association was the most significant association among all the phenotypes assessed, emphasizing the high burden of neurodevelopmental features in *SCN8A*-related disorders. When breaking down more specific neurodevelopmental features, developmental delay was significantly more common in the *SCN8A* cohort between 4 and 12 years of age. More specific phenotypes enriched in the *SCN8A* cohort included delayed fine motor development, expressive language delay, motor delay, persistent head lag, and receptive language delay (**Supplementary Table 2**). Among these more specific neurodevelopmental features, receptive language delay showed the highest significance at 4.5 (*P* = 8.68 × 10^−4^, OR 27.51, CI 3.59-210.82) and 5 (*P* = 1.61 × 10^−4^, OR 86.22, CI 6.76-4431.54) years of age, indicating an over 20-fold enrichment. No neurodevelopmental phenotypes were less likely to occur in the *SCN8A* cohort.

In terms of motor features, hypotonia was more common in the *SCN8A* cohort, which was most prominently observed at 4.25 years when this feature was almost ten times more likely than in other epilepsies with known or presumed genetic etiologies (*P* = 1.49 × 10^−14^, OR 9.96, CI 5.30-19.36). Abnormal muscle tone, a more general clinical feature was only five times more likely in *SCN8A*-related disorders at the same age, emphasizing the specificity of hypotonia in individuals with *SCN8A*-related disorders (*P* = 8.84 × 10^−9^, OR 5.43, CI 2.89-10.63). In contrast, limb hypertonia in infancy was almost 40 times more probable to occur in the *SCN8A* cohort compared to other epilepsies (4 months of age, *P* = 8.16 × 10^−8^, OR 37.74, CI 8.73-227.55).

We also identified other clinical features related to motor function that were significant after multiple testing. Tone-related features included axial hypotonia, facial hypotonia, spastic tetraplegia, and lower limb hypertonia, emphasizing the high frequency of muscle tone abnormalities in *SCN8A*-related disorders. Even though we observed an increased enrichment for limb hypertonia, individuals with *SCN8A*-related disorders were 5-10 times less likely to present with spasticity and spastic diplegia, an association most prominent at one year (*P* = 1.00 × 10^−6^, OR 0.11, CI 0.02-0.36) and 1.25 years (*P* = 1.60 × 10^−6^, OR 0.09, CI 0.01-0.34), respectively (**Supplementary Table 2**).

### *SCN8A*-related disorders phenotypes cluster within clinically relevant subgroups

To delineate subgroups within *SCN8A*-related disorders, we examined how clinical features progress over time for individuals depending on the functional consequence of the variant and for specific variant hotspots and recurrent variants (**Figure 3**). Compared to the remainder of the broader *SCN8A* cohort, individuals with established GOF *SCN8A* variants were more likely to present with global developmental delay and seizures, especially in the first year of life. In particular, the association with generalized-onset seizures was the strongest at nine months (*P* = 0.005, OR 3.85, CI 1.35-11.55) and with global developmental delay at three months (*P* = 0.002, OR 5.67, CI 1.74-20.23). Other clinical features nominally significant for the GOF subgroup included abnormal muscle tone, including hypotonia and axial hypotonia, and an up to 7-times higher frequency of various seizure types including focal-onset seizures, motor seizures, and tonic seizures (**Supplementary Table 3**). In contrast, clinical features more common in individuals with established LOF *SCN8A* variants appeared later in childhood after the age of four, including atypical absence seizures, which were more prominent at 4.25 years (*P* = 0.013, OR 32.71, CI 1.44-2193.51), delayed speech and language development, which were more than 30 times more likely at 7.75 years (*P* = 0.013, OR 32.71, CI 1.44-2193.51), and global developmental delay, which was most significantly associated in *SCN8A* LOF at 8.5 years (*P* = 0.029, OR 7.26, CI 0.95-87.21). All associations with GOF and LOF subgroups were only nominally significant.

**Figure 3.**
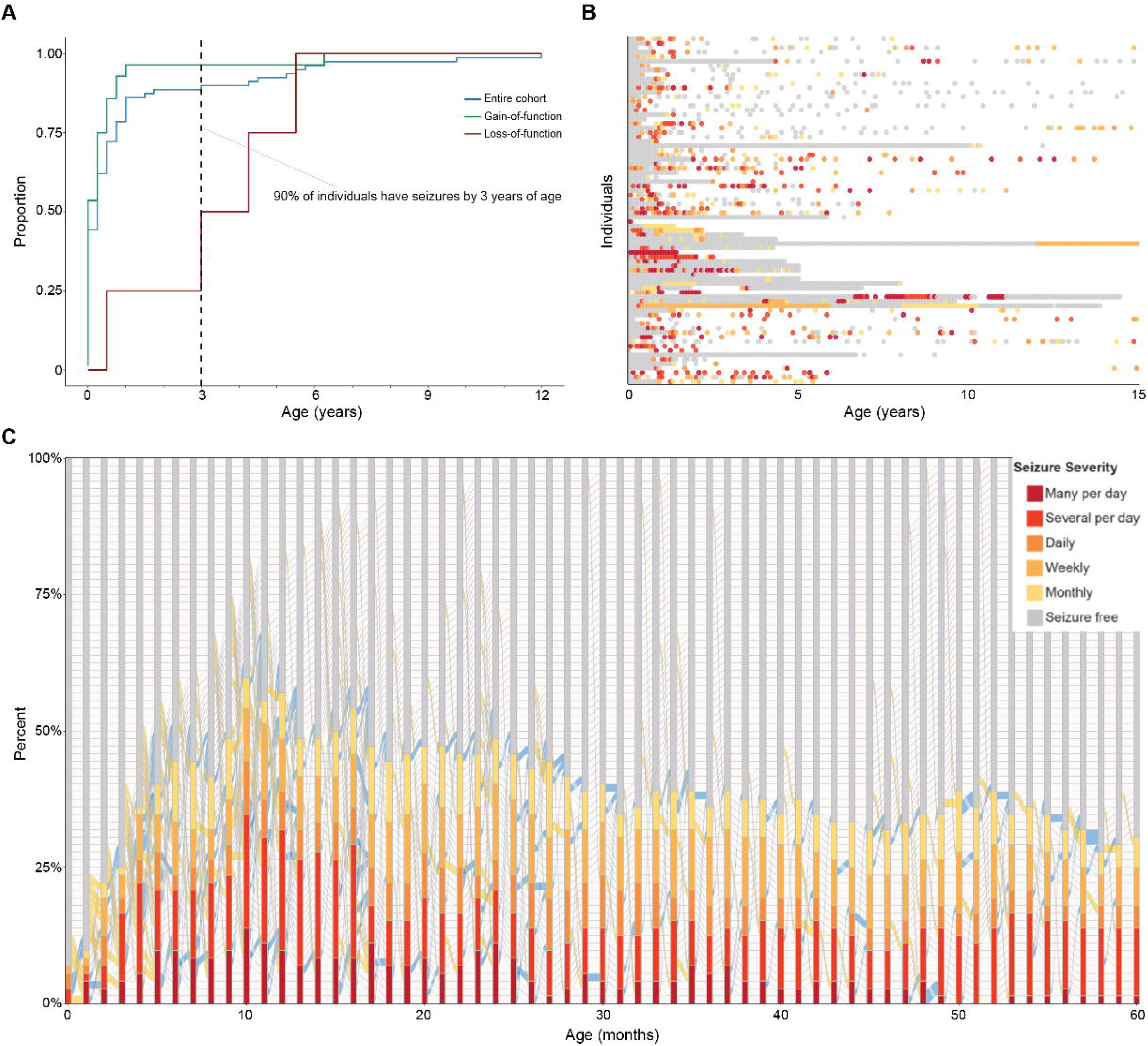
The seizure landscape of *SCN8A*-related disorders. Seizure onset in individuals with *SCN8A*-related disorders (blue – entire cohort, green – GOF sub-cohort, red – LOF sub-cohort) **(A)**. Seizure severity over time on a 0-5 scale from seizure freedom (SF = 0) to more than 5 seizures per day (SF = 5) **(B)**. Seizure history reconstruction in monthly increments in the first five years of life (blue connecting lines - seizure frequency reduction, yellow connecting lines - seizure frequency increase, grey connecting lines - no change in seizure frequency) **(C)**.

We applied a comparable longitudinal phenotypic analysis for individuals carrying recurrent *SCN8A* variants compared to the wider *SCN8A* cohort. For those with the p.Arg1872Trp/Gln/Leu hotspot variant (*n*=6), the most prominent age-related phenotypes included limb hypertonia most prominent at 3 months (*P* = 5.52 × 10^−4^, OR 31.93,CI 3.49-453.54), generalized-onset seizures which were more than ten times more likely at 6 months (*P* = 0.011, OR 12.42, CI 1.28-616.77), and generalized hypotonia, which was almost ten times more likely as early as three months (*P* = 0.015, OR 9.30, CI 1.19-113.08); other nominally significant associations for the p.Arg1872Trp/Gln/Leu hotspot variant included axial hypotonia, neonatal seizure, and bilateral tonic-clonic seizure (**Supplementary Table 4**).

Individuals carrying the recurrent p.Arg850Gln (*n*=5) variant were more than 40 times more likely to present with focal motor seizure at birth (*P* = 0.009, OR 42.53, CI 1.80-2936.94), myoclonic seizure at 6 months (*P* = 0.009, OR 41.96, CI 1.78-2899.93) and tonic seizures most prominent at 2.25 years (*P* = 0.009, OR 42.53, CI 1.80-2936.94), and myoclonic seizure at 0.5 years (*P* = 0.009, OR 41.96, CI 1.78-2899.93). Apart from these, phenotypes that had nominal significance included persistent head lag, motor seizures, infantile spasms, hypotonia, neurodevelopmental delay, limb hypertonia, and focal-onset seizures.

Six individuals with the recurrent p.Arg1617Gln variant had a high enrichment global developmental delay peaking at six months (*P* = 0.014, OR Inf, CI 1.25-Inf), followed by hypotonia that became most prominent at 3.5 years (*P* = 0.007, OR Inf, CI 1.55-Inf). Other nominally significant phenotypes for carriers of the recurrent p.Arg1617Gln variant included the more general feature of abnormal muscle tone, as well as focal-onset seizures and generalized-onset seizures.

Individuals with recurrent p.Gly1475Arg variant (*n*=4) had a high frequency of seizures later during childhood, with the strongest association seen at 11.75 years (*P* = 0.001, OR Inf, CI 3.10-Inf). In contrast to the prominence of seizures in the second decade of life, we also observed a higher frequency of other neurological and neurodevelopmental features in the first three years of life in carriers of the recurrent p.Gly1475Arg variant, including delayed gross motor development that was more than 15 times more likely at 1.25 years (*P* = 0.025, OR 16.84, CI 0.99-291.58), parallelled by a strong association with the more general clinical concept of motor delay, which peaked later at 2.25 years with a more than 10 times higher likelihood of occurrence (*P* = 0.035, OR 13.52, CI 0.83-224.36). Other features nominally associated with the recurrent p.Gly1475Arg variant were abnormal muscle tone including hypertonia and axial hypotonia, as well as global developmental delay, delayed speech and language development, and bilateral tonic-clonic seizures.

### The longitudinal *SCN8A* epilepsy landscape is dynamic

Assessing the overall epilepsy landscape of *SCN8A*-related disorders independent of age-related features, we found that the most common seizure phenotypes were generalized-onset seizure (*f* = 0.91), motor seizure (*f* = 0.75), focal-onset seizure (*f* = 0.65), bilateral tonic-clonic seizure (*f* = 0.61), epileptic/infantile spasm (*f* = 0.30/0.28), tonic seizure (*f* = 0.30), and non-motor seizure (*f* = 0.29, **Table 2**).

**Table 2.**
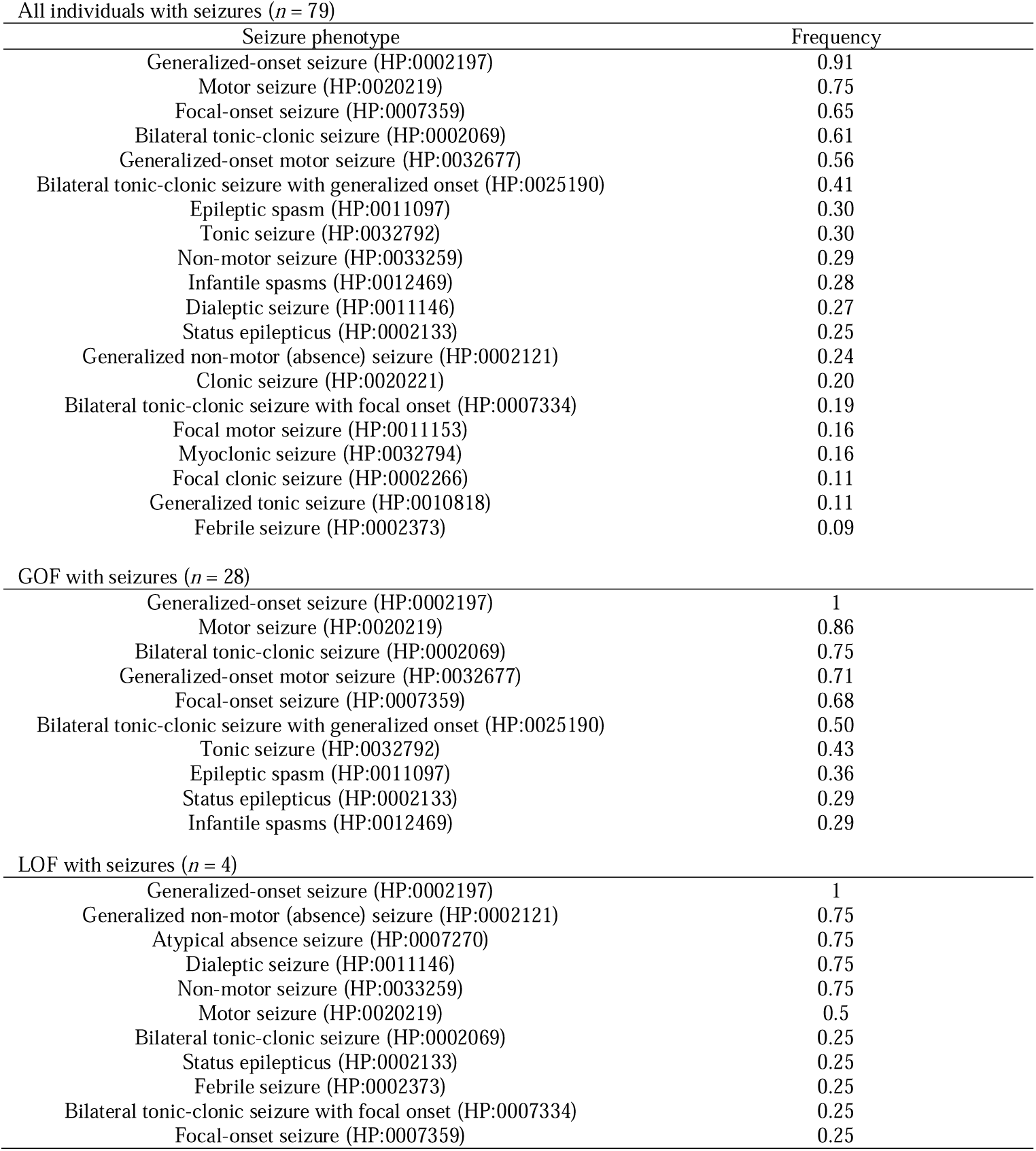
Most common seizure phenotypes.

Seizure onset in individuals with *SCN8A* epilepsy was most prominent in infancy through early childhood. By three months, 50% of the cohort had their first seizure, and 90% of the cohort had their first seizure by three years of age (**Figure 3A**). When analyzing seizure onset depending on the biophysical consequence of the *SCN8A* variant, individuals with GOF variants (mean onset = 0.42 years) had their first seizure earlier than those with LOF variants (mean onset = 3.31 years; *P* = 0.016). The assessment of the monthly longitudinal seizure progression (**Supplementary Figure 1**) revealed a heterogeneous picture with regards to individual seizure frequencies with frequent transition between seizures frequencies, including periods of seizure freedom (**Figure 3B**). Over half of the overall *SCN8A* cohort had seizures at 10 months of life, when the highest seizure frequencies (> 5 per day and 2-5 per day) were also most common. Within the first 10 month of life, the seizure frequencies were most dynamic, exhibiting the greatest number of changes in monthly seizure frequencies.

### *SCN8A*-specific treatment strategies are validated by ASM prescription patterns and analysis of comparative effectiveness

Medication prescription data (**Figure 4A**) showed that the most commonly prescribed ASMs in *SCN8A*-related disorders were levetiracetam (*f* = 0.67), oxcarbazepine (*f* = 0.62), clobazam (*f* = 0.38), phenobarbital (*f* = 0.38), lacosamide (*f* = 0.36), phenytoin (*f* = 0.30), the ketogenic diet (*f* = 0.29), topiramate (*f* = 0.26), lamotrigine (*f* = 0.25), valproate (*f* = 0.23), and cannabidiol (*f* = 0.19). When examining medication prescription relative to the timing of receiving the *SCN8A* diagnosis, we identified four ASMs that were exclusively prescribed only after the genetic diagnosis was made: cannabidiol, rufinamide, perampanel, and gabapentin (**Figure 4B**).

**Figure 4.**
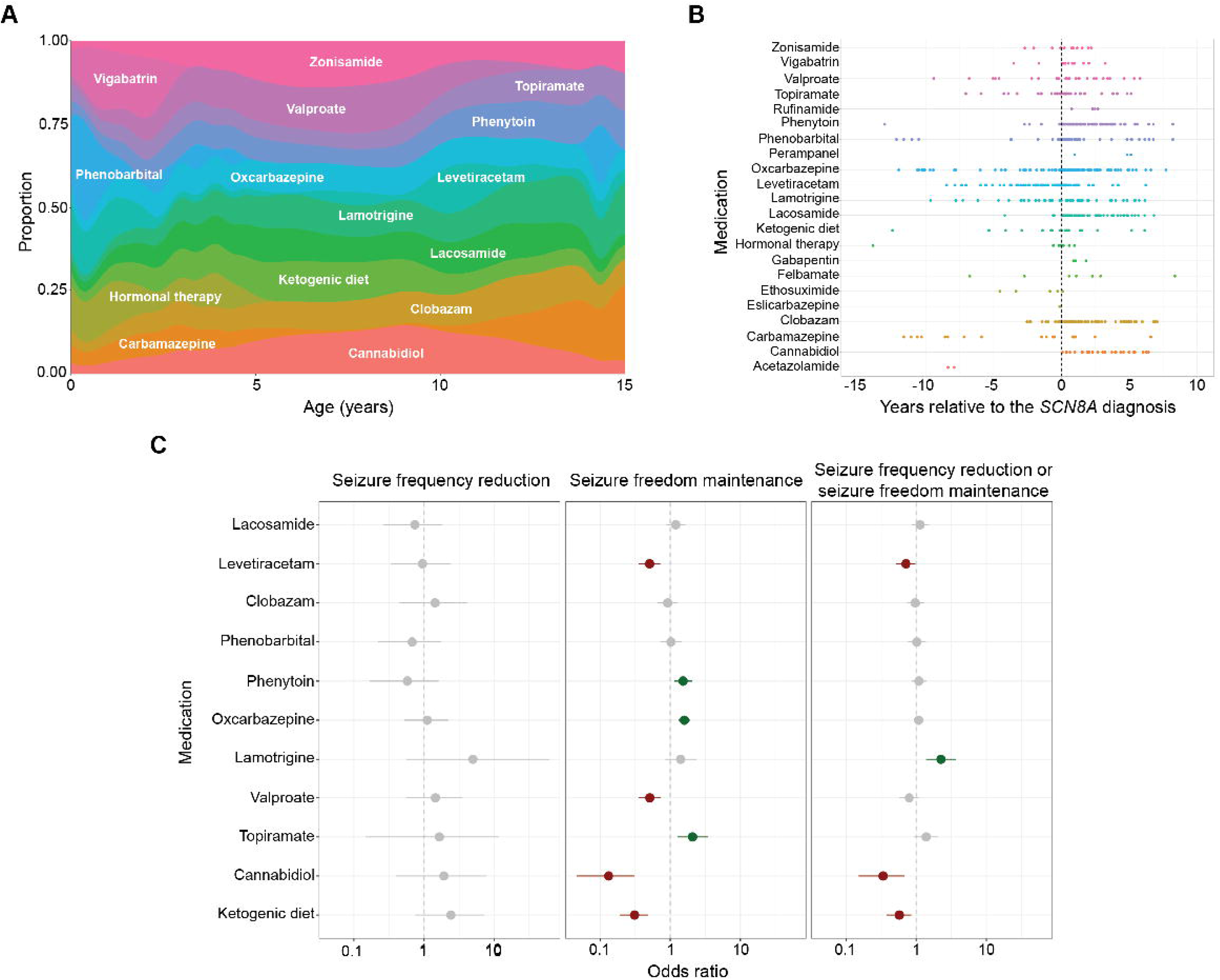
Medication data. Medication prescription landscape **(A)**. Prescription patterns before and after the *SCN8A* diagnosis **(B)**. Medication effectiveness analysis (green – nominally significant association (P < 0.05 with OR > 1, grey – non-significant association, red – nominally significant association (P < 0.05 with OR < 1) **(C)**.

We next performed a joint analysis to assess which ASMs were effective in reducing seizure frequency and maintaining seizure freedom (**Figure 4C**). No single ASM demonstrated significant effectiveness in decreasing seizure frequencies alone. However, various ASMs were associated with maintaining seizure freedom in individuals with *SCN8A*-related disorders (**Figure 4C**), including oxcarbazepine (*P* = 7.22 × 10-7, OR 1.59, CI 1.32-1.92), phenytoin (*P* = 0.004, OR 1.53, CI 1.14-2.05), and topiramate (*P* = 0.003, OR 2.08, CI 1.27-3.49). When assessing the ability of each ASM to reduce seizure frequency or maintain seizure freedom, lamotrigine (*P* = 5.21 × 10-4, OR 2.26, CI 1.39-3.72) was the only ASM demonstrating increased comparative effectiveness.

### Developmental outcomes

From the available developmental outcome data (**Figure 5**), we determined that 30 out of 54 individuals were able to achieve independent ambulation (median = 1.92 years old, interquartile range [IQR] 1.44-2.50 years old), 34 out of 47 were able to communicate using spoken language (defined as using at least a single word, median = 1.44 years old, IQR 1.01-3.07 years old), 41 out of 52 were able to reach (median = 8.2 months old, IQR 0.39-1.33 years old), 20 out of 46 were able to control head posture (median = 7.7 months old, IQR 0.42-1.73 years old), 30 out of 44 were able to roll (median = 6.7 months old, IQR 0.36-0.93 years old), 24 out of 49 were able to sit unsupported (median = 10.8 months old, IQR 0.68-1.51 years old), 27 out of 41 were able to grasp (median = 7.2 months old, IQR 0.38-1.01 years old), and 20 out of 35 were able to follow commands (median = 3.22 years old, IQR 2.32-5.70 years old); not all individuals were assessed for all measures. This analysis demonstrates that development in *SCN8A*-related disorders can be quantified, allowing for prediction of milestone acquisition in this patient cohort.

**Figure 5.**
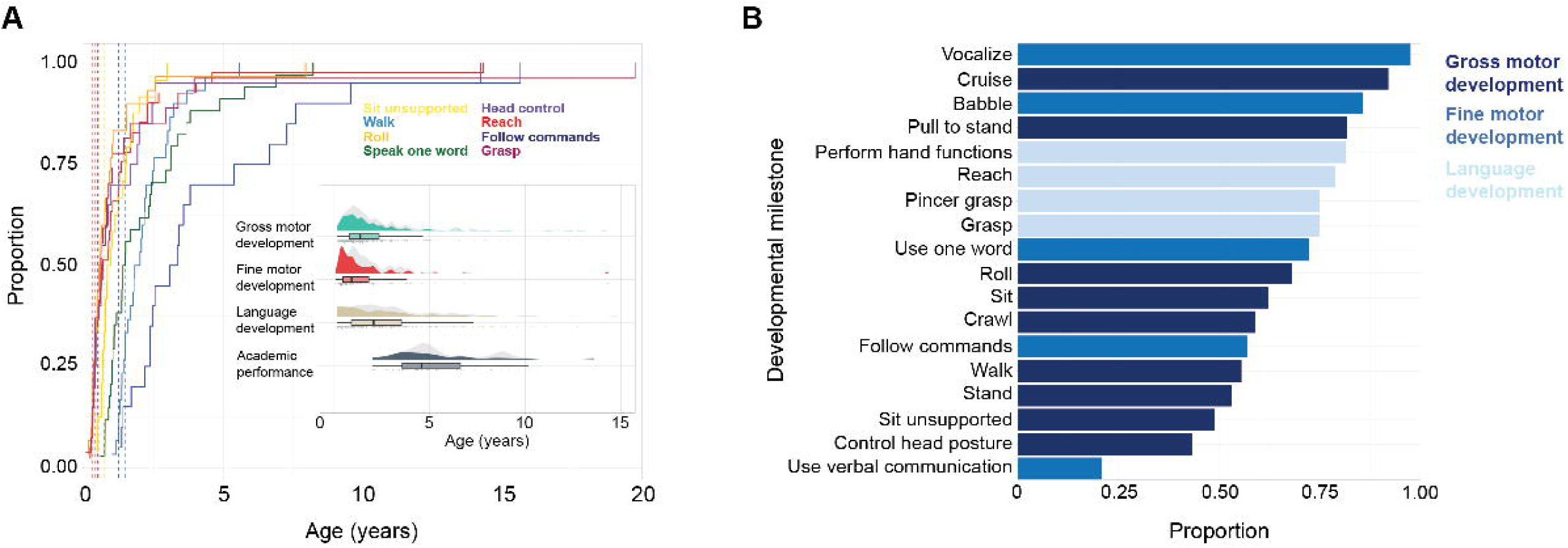
Developmental milestones. Cumulative onset of developmental milestone acquisition (light blue – walk, green – one word, red – reach, purple – head control, orange – roll, yellow – sit unsupported, maroon – grasp, blue – follow commands) **(A)**. Proportion of the achieved developmental milestones **(B)**.

## Discussion

Leveraging large-scale EMR data, we comprehensively characterized the longitudinal clinical landscape of *SCN8A*-related disorders in 82 individuals across 604 patient years. Through this analysis, we identified four main findings. First, *SCN8A*-related disorders stand out from a broader range of genetic epilepsies by the high frequency of generalized-onset and bilateral tonic-clonic seizures in infancy. Second, analyzing clinical patterns linked to biophysical consequences and recurrent variants revealed unique longitudinal trajectories, emphasizing the inherent heterogeneity within *SCN8A*-related disorders. Third, we found real-world evidence that sodium channel blockers including oxcarbazepine, phenytoin, lamotrigine, and topiramate show superior effectiveness in *SCN8A*-related disorders to maintain seizure freedom. Fourth, we were able to establish that *SCN8A*-related disorders follow a unique trajectory of developmental milestone acquisitions, allowing for more precise prognoses, and first steps towards clinical trial readiness by establishing reliable outcomes measures.

The use of electronic medical record data and particularly real-world data has only been established within the last few years.^16^ Accordingly, demonstrating the validity of results emerging from the electronic medical record analyses is critical. Our findings regarding the general profile of *SCN8A*-related disorders are consistent with prior reports. In particular, we find that *SCN8A*-related disorders have prominent clinical features starting early in life, including a high frequency of seizures as well as neurodevelopmental, and muscle tone abnormalities.^2,5,33,34^ While generalized-onset seizures are associated with multiple genetic epilepsies, including *SCN1A*-, *SYNGAP1*-, *CHD2*-, and *SLC6A1*-related disorders,^35–38^ the early age of seizure onset and the early emergence of bilateral tonic-clonic seizures is a feature strongly associated with *SCN8A*-related disorders, which have an almost 50-fold increased risk of this seizure type in late infancy. The high frequency of bilateral tonic-clonic seizures contributes to the morbidity and potentially mortality given the secondary autonomic features and post-ictal electrographic suppression associated with this seizure type, both of which are markers of sudden unexpected death in epilepsy (SUDEP) risk.^39–43^ The overall neurodevelopmental features of *SCN8A*-related disorders reflect those of a developmental and epileptic encephalopathy: neurodevelopmental delay is common in *SCN8A*-related disorders and almost 2 times more likely after 3 months of life compared to a wide range of known or presumed genetic epilepsies. Furthermore, abnormalities in muscle tone, including both hypotonia and hypertonia, were more common among those with *SCN8A*. Importantly, the most prominent phenotypic associations with *SCN8A*-related disorders were seen in the first five years of life, highlighting the early morbidity in this condition compared to other epilepsies.

Assessing longitudinal genotype-phenotype correlations in *SCN8A*-related disorders, we established clinically important differences in those carrying GOF, LOF, and recurrent variants. In particular, individuals with known GOF variants distinguish themselves from the remainder of the *SCN8A* cohort by increased frequency of generalized-onset seizures, global developmental delay, and abnormal muscle tone manifesting in the first months of life. Conversely, those with LOF variants exhibited absence seizures and a strong developmental phenotype emerging by 5 years of age.

Genotype-phenotype correlations in *SCN8A*-related disorders extend beyond loss-versus gain-of-function biophysical consequences. Assessing clinical features of individuals with recurrent variants revealed specific age-related unique phenotypes. While all the recurrent variants we analyzed were classified as GOF, there were distinct patterns in observed phenotypes and the timing of their occurrence. Through this analysis, we were able to identify novel clinically relevant genotype-phenotype associations, such as p.Arg850Gln-epileptic spasms, p.Arg1872Trp/Gln/Leu-neonatal seizures, and p.Arg1617Gln-focal seizures. Interestingly, those carrying p.Gly1475Arg had the most significant phenotypic associations after 7 years. In contrast, all three other recurrent variants showed the most prominent genotype-phenotype associations within the first 4 years of life.

Combining the monthly reconstruction of seizure frequencies and medication prescription data, we identified oxcarbazepine, phenytoin, lamotrigine, and topiramate as effective in seizure management in *SCN8A*-related epilepsy. Our study provides real-world evidence to a conceptual framework that had previously only been established in translational research studies and small case series, namely the effectiveness of sodium channel blockers in *SCN8A*-related epilepsy given the predominant GOF mechanism in this channelopathy.^2,7,44^ Importantly, no single anti-seizure medication showed superior effectiveness in reducing seizure frequencies, emphasizing the refractory and sometimes erratic nature of *SCN8A*-related seizures, particular in the first years of life. However, our analysis demonstrated that if seizure freedom was achieved, its maintenance was associated with the use of sodium channel blockers such as oxcarbazepine, phenytoin, or topiramate. The medication effectiveness analysis was conducted for all individuals in the cohort, not only individuals carrying GOF variants, suggesting that those whose variants have not been functionally studied on the electrophysiological level to date could benefit from a sodium channel blocker regimen, even though not all individuals may benefit given the specific biophysical abnormalities linked to specific variants. Of note, cannabidiol was an agent prescribed only after *SCN8A* genetic diagnosis; however, our analysis demonstrated that the effectiveness of cannabidiol was limited. It was neither associated with seizure freedom maintenance nor with seizure frequency reduction.

Lastly, the analysis of the developmental outcome data for individuals with *SCN8A*-related disorders underscore the significant developmental consequences of this condition. Neurodevelopmental features in *SCN8A*-related disorders can be observed across multiple domains, with a particularly limited acquisition of gross motor skills. The pattern of developmental outcome attainment in *SCN8A*-related disorders resembles that seen in *STXBP1*-related disorders, which have been analyzed using a similar framework previously.^24^ This suggests that *SCN8A*-related disorders may follow a more global developmental trajectory common across the genetic developmental and epileptic encephalopathies, potentially allowing for joint outcome measures across various genetic epilepsies.

In summary, our study represents a first longitudinal reconstruction of the natural history of *SCN8A*-related disorders, one of the most common genetic epilepsy-related channelopathies. Through a standardized approach to retrospective phenotypic data documented in the EMR, we identified clinically relevant features that can serve as a launching point for more in-depth prospective clinical trial readiness studies to develop standardized outcome measures in *SCN8A*-related disorders.

## Acknowledgments

J.L.M. was supported by the CURE Epilepsy Rare Epilepsy Partnership Award for SCN8A, co-funded by The Cute Syndrome Foundation.

I.H. was supported by the National Institute of Neurological Disorders and Stroke (R01 NS127830-01A1, R01 NS131512-01 and K02 NS112600).

This work was also supported by intramural funds of the Children’s Hospital of Philadelphia through the Epilepsy NeuroGenetics Inititative (ENGIN).

